# Beyond six feet: The collective behavior of social distancing

**DOI:** 10.1101/2023.10.16.23297077

**Authors:** Zhijun Wu

## Abstract

In a severe epidemic such as the COVID-19 pandemic, social distancing can be a vital tool to stop the spread of the disease and save lives. However, social distancing may induce profound negative social/economic impacts as well. How to optimize social distancing is a serious social, political, as well as public health issue yet to be resolved. This work investigates social distancing with a focus on how every individual reacts to an epidemic, what role he/she plays in social distancing, and how every individual’s decision contributes to the action of the population and vice versa. Social distancing is thus modeled as a population game, where every individual makes decision on how to participate in a set of social activities, some with higher frequencies while others lower or completely avoided, to minimize his/her social contacts with least possible social/economic costs. An optimal distancing strategy is then obtained when the game reaches an equilibrium. The game is simulated with various realistic restraints including (i) when the population is distributed over a social network, and the decision of each individual is made through the interactions with his/her social neighbors; (ii) when the individuals in different social groups such as children vs. adults or the vaccinated vs. unprotected have different distancing preferences; (iii) when leadership plays a role in decision making, with a few leaders making decisions while the rest of the population just follow. The simulation results show how the distancing game is played out in each of these scenarios, reveal the conflicting yet cooperative nature of social distancing, and shed lights on a self-organizing, bottom-up perspective of distancing practices.

## Introduction

In a severe epidemic such as the COVID-19 pandemic, social distancing can be a vital tool to stop the spread of the disease and save lives [1–13]. This is true before a vaccine becomes available and even after the population achieves certain herd immunity, for the pathogen always evolves, and the epidemic may develop into an endemic [14–22].

In general, social distancing is advocated as for people to keep certain distances from each other to reduce close social contacts [23–26]. However, in practice, it means a lot more and involves managing multiple social activities to avoid some of them but keep some others, in order to slow down the spread of infection while containing the negative social/economic impacts [27–51]. It is not clear how to optimize the activities though, often causing confusions and even rejections [52–58].

Some social activities have low contact rates but are socially isolating such as hiking, gardening, staying home, or reading. Some others are socially more involved but have close social contacts such as large gatherings, going to night clubs, going to shopping malls, or watching sports. In between, there are social activities that are essential to our daily life such as grocery shopping, cafeteria dinning, visiting friends, taking buses, or going to schools or workplaces. Given a set of social activities, an individual needs to make decision for how to participate in each of them, some probably with higher frequencies while others lower or even completely avoided, so that his/her close social contacts can be minimized at a least possible social/economic cost.

The decision of each individual must depend on or be influenced by the actions of all other individuals in the population. For example, if everybody decides to stay home, an individual may choose to watch a movie although the risk of having close contacts at a movie theater is high. On the other hand, if the whole population decides to go hiking, he/she may want to avoid it although hiking is usually a low contact activity. Collectively, social distancing can thus be considered as a population game [59, 60], where based on what the population does, every individual makes his/her own decision on how to participate in a given set of social activities so that he/she can minimize his/her social contacts and possible social/economic costs. An optimal distancing strategy can then be obtained when the game reaches an equilibrium.

Research on social distancing has surged since the outbreak of the COVID-19 pandemic, providing a wealth of knowledge and experience on non-pharmaceutical measures for preventing epidemics from spreading. However, most of these studies are about the influences of public policies, economic concerns, or cultural differences on social distancing, but not particularly about social distancing as a human behavior or the collective behavior of a population for that matter. Work on modeling social distancing has been pursued in the past [61–64] including some done recently [65–78], but the focus is on the dynamics of epidemics with changing patterns or levels of social distancing, with little specifics on how the distancing activities are carried out and how certain distancing patterns or levels are achieved.

The work in this paper follows a game theoretic approach to social behavior in general [79–84] and to social distancing in particular [85–88], and investigates social distancing with a focus on how every individual reacts to an epidemic, what role he/she plays in social distancing, and how the individual decision contributes to the action of the population and vice versa. The collective behavior of social distancing is modeled as a population game, where every individual makes a distancing decision and together the population reaches an equilibrium when every individual achieves his/her distancing goal.

A number of issues rise immediately for the general game model for social distancing: First, a general population game assumes that every individual interacts with all others in the population and knows their strategies, which is not true in the real world, where people usually interact only with their social acquaintances. The game also assumes that the individuals are all the same when evaluating the distancing risks and making their distancing decisions, but in reality, they are not. For example, children and adults seem to have different infection rates for COVID-19 and would therefore consider the distancing risks of social activities differently; and so do the vaccinated and unprotected individuals. The general game also requires every individual to be able to make rational decisions for the game to eventually reach equilibrium. The condition is again unrealistic, for not everyone is able to or willing to make his/her own decisions.

However, all these issues can be addressed by making several refinements on the general model: First, the population can be assumed to be distributed over a social network, and the decision of each individual can be made through the interactions with his/her social neighbors. Such a network can be simulated by generating a small-world network using for example the Watts-Strogatz algorithm [89]. The simulation results presented in this paper show that the distancing game can be played successfully on such a network. Surprisingly, the game approaches to an equilibrium state as in the general case even when the interactions among the individuals are restricted only to their close neighbors.

Second, the population can be divided into different social groups according to certain social/biological/medical characteristics such as the age of the individuals (e.g., children, adults, seniors, etc.) or the level of protection (e.g., the vaccinated, recovered, unprotected, etc.) or the economic vulnerability (income above average, middle income, low income, etc.). The distancing risks of participating certain social activities can then be evaluated using different criteria for different social groups. Theoretical and simulation results for the distancing game in such heterogeneous populations are discussed in the paper, showing that the game can be played in almost the same form as in the general case: just use different risk-assessment functions for different social groups; and if the interactions among the individuals are restricted within their own groups, each social group would eventually find its own equilibrium strategy while the whole population approaches to the average one.

Third, a small number of individuals can be selected to act as leaders and the rest of the population as followers. The leaders make decisions on their own strategies while the followers simply copy the strategies of the leaders. The distancing game can then be carried out with such a leader-follower scheme. The simulation results show that the game can indeed proceed without requiring every individual to make rational decisions, and reach its equilibrium successfully even when only a small number of individuals, say 20% of the population, are designated as leaders. Indeed, in practice, it is most likely that only a small number of individuals such as public health experts or community leaders make decisions or recommendations while all others just follow.

As such, the work in this paper confirms the game model as a plausible approach to the study of the collective behavior of social distancing. It shows how the distancing game is played out in possible realistic as well as idealistic scenarios, reveals the conflicting yet cooperative nature of social distancing, and sheds lights on a self-organizing, bottom-up perspective of distancing practices.

## Results

### As a population game

Consider a population of *m* individuals with *n* social activities. Assume that every individual needs to decide a frequency to participate in each of the activities, say *x*_*i*_ for activity *i*. Then the collection of these frequencies *x* = {*x*_*i*_ : *i* = 1, …, *n*} can be considered as a distancing strategy of the individual. Let *y*_*i*_ be the average frequency of the population to participate in activity *i*. Then, the collection of these average frequencies *y* = {*y*_*i*_ : *i* = 1, …, *n*} can be considered as a distancing strategy of the population. Here a frequency *x*_*i*_ or *y*_*i*_ can be represented by the active hours in activity *i* in a week (total 112 hours per week if 16 hours are counted as active hours per day excluding 8 hours sleeping time).

Given a distancing strategy *y* of the population, assume that the potential distancing risk of having close social contacts and negative social/economic impacts in activity *i*, when fully participated, can be represented by a function *p*_*i*_(*y*). Then, the distancing risk of an individual of strategy *x* at activity *i* must be *x*_*i*_*p*_*i*_(*y*), and at all the activities together be Σ_*i*_ *x*_*i*_*p*_*i*_(*y*), where Σ_*i*_ means the sum over all *i*’s. Let this summation be denoted as a function *π*(*x, y*). A distancing game can then be defined for an individual against the population with *π*(*x, y*) as the cost function; and a strategy *x*^∗^ is an equilibrium strategy for the game if and only if every individual in the population takes this strategy (and hence *y*^∗^ = *x*^∗^), and his/her distancing risk *π*(*x*^∗^, *y*^∗^) using strategy *x*^∗^ is no greater than the distancing risk *π*(*x, y*^∗^) using any other strategy *x*.

To make the matter simpler, assume that the social activities are independent of each other, i.e., the individuals participating in one of the activities do not have contacts with those in other activities. When the activities are independent, the function *p*_*i*_(*y*) can be defined to have two parts, one for close social contacts and the other for negative social/economic impacts, both proportional only to the participating frequency *y*_*i*_ of the population in activity *i*. The amount of close social contacts can be estimated by a term *α*_*i*_*y*_*i*_ with *α*_*i*_ named as the contact factor of activity *i*. The amount of negative social/economic impacts can be estimated by a term *β*_*i*_*y*_*i*_ with *β*_*i*_ named as the impact factor of activity *i*. The unit for *β*_*i*_ is the same as *α*_*i*_, i.e., the number of contacts per hour, but for *α*_*i*_ it is the number of close contacts one may have if fully participating in activity *i*, while for *β*_*i*_ it is the number of necessary contacts one may lose when staying only in this activity. Table 1 gives a list of *α*_*i*_ and *β*_*i*_ values assigned to 20 commonly attended social activities or CASA activities for short. They are estimated based on common practices or public guidelines such as the CDC social distancing recommendations [26] (details in **Deriving contact and impact factors** in **Methods**). They will be used throughout the paper for the purpose of concept proofing. In real applications, they certainly need to be further refined and justified with more experimental data.

**Table 1.** Estimated contact factors and impact factors for 20 CASA activities.

| Act: | 1 | 2 | 3 | 4 | 5 | 6 | 7 | 8 | 9 | 10 |
| --- | --- | --- | --- | --- | --- | --- | --- | --- | --- | --- |
| $\alpha_i$ : | 1 | 1 | 1 | 1 | 2 | 2 | 2 | 2 | 3.5 | 3.5 |
| $\beta_i$ : | 2.8000 | 2.8000 | 4.6667 | 4.6667 | 2.8000 | 4.6667 | 4.6667 | 2.8000 | 2.8000 | 4.6667 |
| Act: | 11 | 12 | 13 | 14 | 15 | 16 | 17 | 18 | 19 | 20 |
| $\alpha_i$ : | 3.5 | 3.5 | 7 | 7 | 7 | 7 | 14 | 14 | 14 | 14 |
| $\beta_i$ : | 4.6667 | 2.8000 | 3.5000 | 3.5000 | 0.8750 | 0.7000 | 2.3333 | 2.3333 | 3.5000 | 3.5000 |

**Table Legends:** Act – activity: 1 - reading or watching TV, 2 - working at home, 3 - hiking, 4 - gardening, 5 - stay with family, 6 - grocery shopping, 7 - go to hospitals, 8 - visit friends, 9 - cafeteria dinning, 10 - go to shopping malls, 11 - taking buses, 12 - going to church, 13 - watch sports, 14 - attend concerts, 15 - go to schools, 16 - go to workplaces, 17 - large gathering, 18 - go to bars or night clubs, 19 - air traveling, 20 - go to movie theaters; $\alpha_i$ – contact factors; #contacts made per hour; $\beta_i$ – impact factors: #contacts lost per hour

By introducing another parameter *δ*_*i*_, the function *p*_*i*_(*y*) can be defined as *p*_*i*_(*y*) = *δ*_*i*_*α*_*i*_*y*_*i*_ + (1 − *δ*_*i*_)*β*_*i*_*y*_*i*_ = *w*_*i*_*y*_*i*_ with *w*_*i*_ = *δ*_*i*_*α*_*i*_ + (1 − *δ*_*i*_)*β*_*i*_ named as the risk factor of activity *i*. Here, 0 ≤*δ*_*i*_ ≤1 can be used to balance between the risks of having close social contacts and negative social/economic impacts at activity *i*. Based on standard game theory, given *p*_*i*_(*y*) = *w*_*i*_*y*_*i*_, if *w*_*i*_ *>* 0 for all *i*, an equilibrium strategy *x*^∗^ can be obtained with 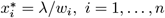, where *λ* is a constant such that 1*/λ* = Σ_*i*_ 1*/w*_*i*_ (more details in **Equilibrium strategies and stabilities** in **Methods**).

Table 2 gives the equilibrium strategies of the distancing game for the 20 CASA activities in Table 1, with *δ*_*i*_ = 1, 0, or 0.5 for all *i* corresponding to the cases for complete social distancing, no social distancing, or something in between, respectively. Fig 1 further illustrates the contrasts among these strategies, where for complete social distancing (blue line, *δ*_*i*_ = 1), for the first 8 activities with small contact factors, the participating frequencies are high, while for the last 8 activities with large contact factors, the participating frequencies are low. On the other hand, if no social distancing (red line, *δ*_*i*_ = 0), for the first 8 activities, the participating frequencies are much lower than those for complete social distancing, while for the last 8 activities, the participating frequencies are much higher. When both social contacts and negative impacts are concerned (brown line, *δ*_*i*_ = 0.5), for the first 8 activities, the participating frequencies are still higher than those with no social distancing, but not as high as those for complete social distancing, and for the last 8 activities, the participating frequencies are certainly lower than those with no social distancing, but not as low as those for complete social distancing.

**Table 2.** Equilibrium strategies for 20 CASA activities w/o social distancing.

| Act: | 1 | 2 | 3 | 4 | 5 | 6 | 7 | 8 | 9 | 10 |
| --- | --- | --- | --- | --- | --- | --- | --- | --- | --- | --- |
| $\delta_i = 1$ : | 14 | 14 | 14 | 14 | 7 | 7 | 7 | 7 | 4 | 4 |
| 0.5 : | 10.370 | 10.370 | 6.9537 | 6.9537 | 8.2092 | 5.9106 | 5.9106 | 8.2092 | 6.2546 | 4.8250 |
| 0 : | 5 | 5 | 3 | 3 | 5 | 3 | 3 | 5 | 5 | 3 |
| Act: | 11 | 12 | 13 | 14 | 15 | 16 | 17 | 18 | 19 | 20 |
| $\delta_i = 1$ : | 4 | 4 | 2 | 2 | 2 | 2 | 1 | 1 | 1 | 1 |
| 0.5 : | 4.8250 | 6.2546 | 3.7528 | 3.7528 | 5.0037 | 5.1174 | 2.4125 | 2.4125 | 2.2517 | 2.2517 |
| 0 : | 3 | 5 | 4 | 4 | 16 | 20 | 6 | 6 | 4 | 4 |

**Fig. 1.**
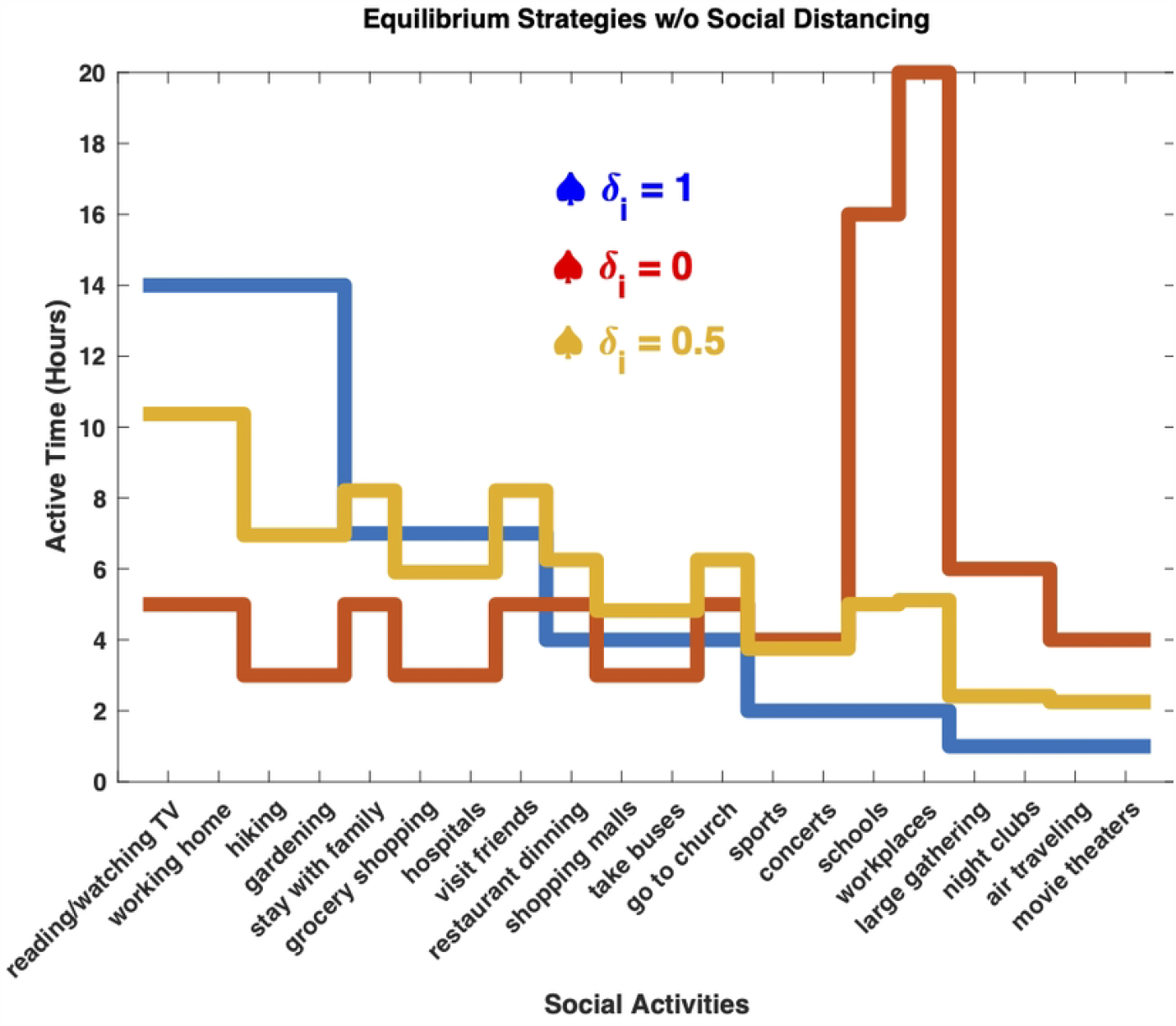
The equilibrium strategies of the distancing game for complete social distancing (blue line), no social distancing (red line), and partially social distancing (brown line), displayed as active times in hours per week (total 112 active hours)

### In small-world social networks

To be more realistic, assume that the population is distributed over a social network, and each individual only interacts with his/her neighbors in the social network. The distancing game can then be viewed as one played by every individual against the population in his/her neighborhood in the social network. Depending on the neighborhood size, the game becomes against a fraction of population ranging from the immediate neighbors of the individual to the whole population. For convenience, define the neighborhood of size *k* of an individual to be one that includes all the neighbors connected to the individual with up to *k* consecutive connections. Then, if *k* is large enough, the neighborhood would include the whole population.

To mimic a real social network, the Watts-Strogatz algorithm [89] is used to generate a small-world social network. Assume that there are 200 individuals in the population (*m* = 200), the average degree of the nodes is 6 (*K* = 6), and 30 percent of all the links for each node come from random connections (*b* = 0.3). Fig 2 shows the generated network and the distribution of the degrees of the nodes. The nodes are displayed around a circle. About 70 percent of the links are along the edges which are not clearly visible in the graph. The rest of the links are more visible in the interior of the circle. In general, as the degree parameter *K* or the randomness parameter *b* increases, the graph becomes denser with more interior links (details in **Generating small-world social networks** in **Methods**).

**Fig. 2.**
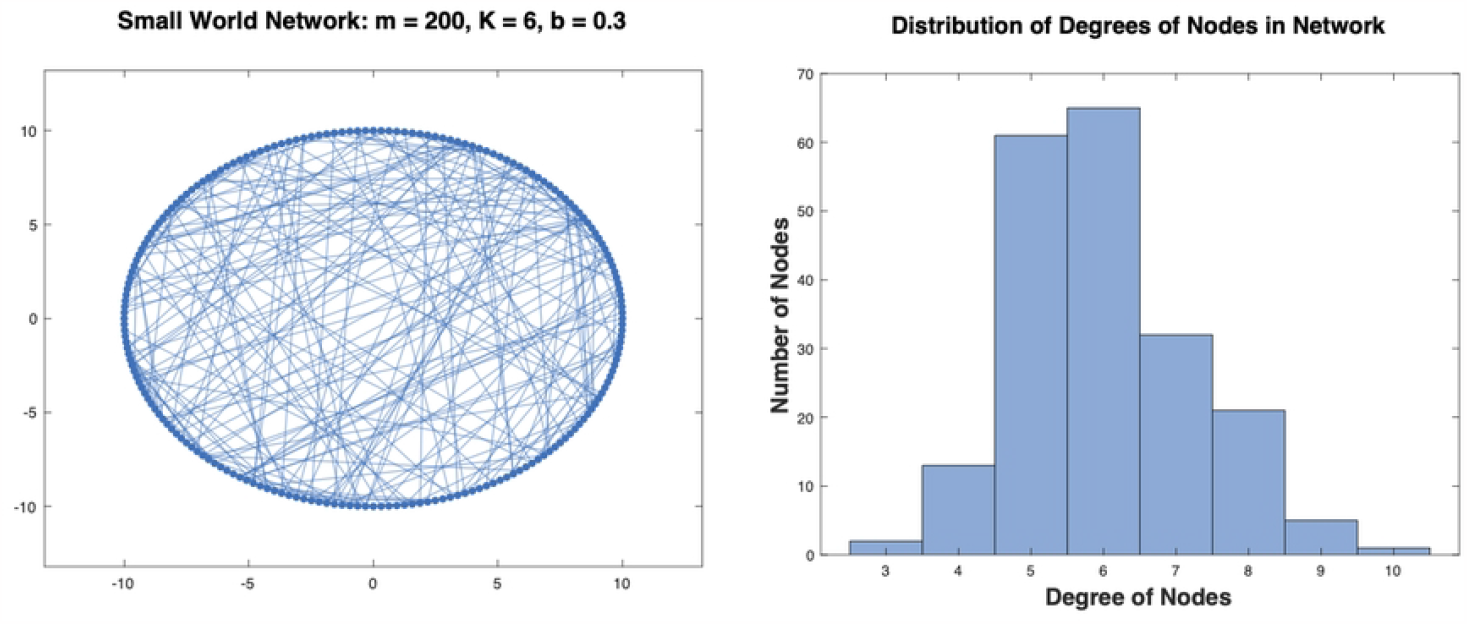
A small-world social network generated by using the Watts-Strogatz algorithm

Assume that every individual plays the distancing game with the population in a neighborhood of the same size. Let *x* be the strategy of any individual and *y* the average strategy in the corresponding neighborhood. Then, the potential distancing risk at activity *i* can still be estimated using the function *p*_*i*_(*y*), with which the individual can update his/her own strategy *x*: If *p*_*i*_(*y*) is higher than the average over all other activities, activity *i* must be riskier, and *x*_*i*_ in activity *i* should be decreased; otherwise be increased. The update can be repeated for every individual in the population until no one can further improve his/her strategy, and the game hopefully reaches an equilibrium.

The distancing game is simulated on networks similar to the one in Fig 2. The simulation algorithm is outlined in **Simulation of distancing games in social networks** in **Methods**. The parameters for the networks are fixed to *m* = 200 and *K* = 6. The activities in Table 1 are used for the game and therefore, *n* = 20. Other parameters for the game are varied with the balancing parameter *δ*_*i*_ = 0, 0.25, 0.5, 0.75, 1 for all *i*, the randomness parameter for the network *b* = 0.10, 0.20, 0.30, 0.40, 0.50, and the neighborhood size *k* = 1, 2, 3, 4, 5, 6. For each set of parameters, an equilibrium strategy *x*^∗^ for every individual and *y*^∗^ for the population for the general distancing game (without network) can be computed directly as described in the previous section. Let *x* be the strategy for an individual obtained by the simulation and *y* the corresponding neighborhood strategy. These quantities are recorded in the simulation and compared with the equilibrium strategies *x*^∗^ and *y*^∗^. The results for all the parameter settings are documented (in Simulation results 1 in Supplementary Information).

Table 3 shows the simulation results for the game on the network in Fig 2, with *δ*_*i*_ = 0, 0.5, 1 respectively for all *i*. The neighborhood size *k* is changed from 1 to 2, 3, 4, 5, 6. Correspondingly, the average number of neighbors for each individual changes from 7 to 28, 92, 179, 199, 200. The last number agrees with the general consensus on small-world social networks where every pairs of individuals can be found connected with up to more or less 6 consecutive connections [90–92]. It is therefore not surprising that when *k* = 6, the game on this network converges to the same equilibrium strategy as the general distancing game (without network), for it is almost the same as the general distancing game played by every individual against the whole population. It is surprising, however, that when the neighborhood size is reduced, even when *k* = 2 with only 28 neighbors in average for each individual, the game still converges to the equilibrium strategy of the general distancing game, with *x* converging to *x*^∗^ and *y* to *y*^∗^. Only for *k* = 1, the game fails to converge, and the simulation terminates when not making any progress. These results are observed to be consistent for all other cases with varying *b* and *δ*_*i*_ values (in Simulation results 1 in Supplementary Information).

**Table 3.** Convergence of distancing strategies in small-world social networks.

| $\delta_i$ | $k =$ | 1 | 2 | 3 | 4 | 5 | 6 |
| --- | --- | --- | --- | --- | --- | --- | --- |
| 0.00 | $\langle \ x - x^*\ \rangle$ | 5.13e-03 | 5.76e-05 | 5.50e-05 | 5.38e-05 | 5.06e-05 | 4.95e-05 |
| | $\langle \ y - y^*\ \rangle$ | 2.71e-03 | 1.16e-05 | 5.51e-06 | 5.20e-06 | 5.89e-06 | 5.56e-06 |
| 0.50 | $\langle \ x - x^*\ \rangle$ | 2.74e-02 | 7.61e-05 | 6.29e-05 | 5.52e-05 | 5.00e-05 | 4.94e-05 |
| | $\langle \ y - y^*\ \rangle$ | 1.04e-03 | 1.49e-05 | 6.52e-06 | 5.93e-06 | 5.31e-06 | 5.44e-06 |
| 1.00 | $\langle \ x - x^*\ \rangle$ | 3.59e-03 | 7.99e-05 | 7.54e-05 | 5.25e-05 | 4.93e-05 | 5.06e-05 |
| | $\langle \ y - y^*\ \rangle$ | 1.97e-03 | 1.64e-05 | 7.21e-06 | 5.77e-06 | 6.49e-06 | 7.31e-06 |

**Table Legends:** $x$ – individual strategy from simulation; $y$ – neighborhood strategy from simulation; $x^*$ – individual strategy at equilibrium (without networks); $y^*$ – population strategy at equilibrium (without network); $\|\cdot\|$ – the Euclidean norm: $\|x\|^2 = x_1^2 + \dots + x_n^2$ ; $\langle \rangle$ – average over all individuals.

### In heterogeneous populations

Not every individual is equally vulnerable for epidemic infection. Nor is every individual equally likely to spread the disease. For example, for COVID-19, children seem not as susceptible to infection as adults, and they are less likely to carry and spread the virus. Similarly, the vaccinated people are more or less immune to infection and are probably more free to participate in social activities than those unvaccinated. Further, the essential workers or economically vulnerable individuals are more concerned with the social/economic impacts than the health benefits of social distancing. A population should therefore be divided into different groups who rate the distancing risks of social activities differently.

As an example, consider a population evenly divided into 4 population groups: either according to the ages of the individuals into *g*_1_: 1-20 years old; *g*_2_: 21-40; *g*_3_: 41-60; and *g*_4_: 61-80, or according to the level of protection of the individuals into *g*_1_: the vaccinated; *g*_2_: the recovered; *g*_3_: the unprotected; and *g*_4_: the most vulnerable. Set *δ*_*i*_ for each of the groups to 0.00 for *g*_1_, 0.25 for *g*_2_, 0.75 for *g*_3_, and 1.00 for *g*_4_, thereby making the first two groups more open to expand their social activities but the last two to prefer more social distancing. Fig 3 demonstrates the results from computer simulation for the distancing game played among these population groups. The simulation is conducted in a similar setting as for the game on the network shown in Fig 2, with the neighborhood size fixed to *k* = 3. In each generation of the simulation, every individual plays the game once, i.e., has a chance to update his/her strategy. However, different from the simulation described in the previous section, when evaluating the potential distancing risks, the contributions from different population groups are different due to their different *δ*_*i*_ values. When they are counted, more weight is also given to the contribution from the individual’s own group than from other groups. In addition, when comparing with the population average on the distancing risks, only the average over the individual’s own group is considered (details in **Distancing in heterogeneous populations** in **Methods**).

**Fig. 3.**
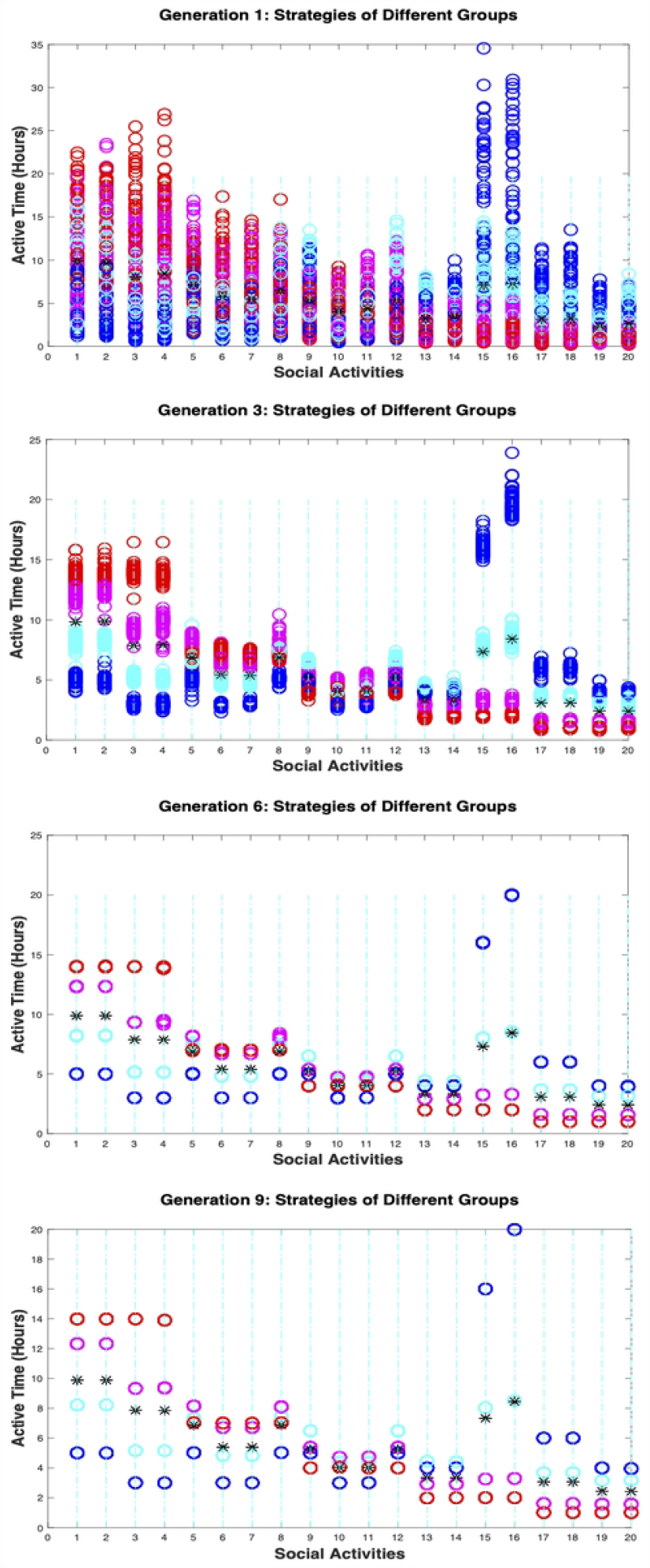
The simulation results for an example distancing game with a heterogeneous population. Shown in the figure are the individual and population strategies of the game in four different generations of the simulation. The stars represent the population strategies and the circles the individual strategies. The circles are color coded for different population groups, with blue for *g*_1_, cyan for *g*_2_, magenta for *g*_3_, and red for *g*_4_. Over each activity, there are 200 circles corresponding to the strategies for 200 individuals. Labeled in the x-axis are 20 CASA activities. Along the y-axis are the active times in hours per week.

Fig 3 displays four snapshots from the simulation, showing the changes of the individual strategies of the game in four different generations. The circles represent the individual strategies and the stars the average population strategies. The circles are color coded for different population groups, with blue for *g*_1_, cyan for *g*_2_, magenta for *g*_3_, and red for *g*_4_. For each activity, there are 200 circles corresponding to the participating frequencies of 200 individuals for the activity. The labels on the *x*-axis are 20 CASA activities as defined in Table 1. The first graph in the figure shows the strategies of the individuals at the beginning of the 1st generation of the simulation, which are randomly generated around reasonably guessed starting strategies for each of the population groups. The second graph shows the strategies of the individuals after the 3rd generation, when they start separating into different groups. The third graph shows the strategies after the 6th generation, when they almost converge to their equilibrium positions. The last graph shows the strategies after the 9th generation, when they are close enough to their equilibrium values, and the simulation is terminated.

In the end of the simulation, each population group reaches an equilibrium strategy or more rigorously, its approximation. Since *δ*_*i*_ = 0.00 and 0.25 for all *i* for *g*_1_ and *g*_2_, the individuals in these two groups are considered to be more risk-taking, and their frequencies to participate in the socially active though high-contact activities (13-20) appear to be higher than the population average, while their frequencies to stay with the low-contact but socially isolating activities (1-8) are lower than the population average. On the other hand, since *δ*_*i*_ = 0.75 and 1.00 for all *i* for *g*_3_ and *g*_4_, the individuals in these two groups are considered to be more conservative, and their frequencies to stay with the low-contact though socially isolating activities (1-8) appear to be higher than the population average, while their frequencies to join the socially active but high-contact activities (13-20) are lower than the population average.

Simulations for distancing games with different population groups are conducted with varying balancing parameter *δ*_*i*_, randomness parameter *b*, and neighborhood size *k*. The results from these simulations (documented in Simulation results 2 in Supplementary Information) are all consistent with what are observed in the above example, showing that the game model can be extended to heterogeneous populations to predict different distancing behaviors among different population groups. What unexpected in these results is that the game for each population group converges to its own equilibrium strategy as if it is played alone, while the population strategy is simply a collective result of all these individual group strategies, which can also be justified in theory (details in **Distancing in heterogeneous populations** in **Methods**). This may not be surprising, on the other hand, as observed during the COVID-19 pandemic that children and adults do have their own distancing strategies as their shares in the average distancing strategy of the population.

### By following the leaders

Not every individual actively participates in social distancing. Even if he/she does, he/she may not necessarily make the decisions as accurately as assumed such as evaluating the distancing risks of the activities and responding with appropriate actions, etc. In practice, it is most likely that only a small number of individuals such as public health experts or community leaders make decisions or recommendations while all others just follow. Indeed, leadership plays an important role in collective actions in both nature and human societies [93–97].

In order to incorporate the leadership factor into the distancing activities, a small number of individuals are designated as leaders and the rest of the population as followers. A leader makes a distancing decision as a regular player in the distancing game, while a follower just copies the strategies of some leaders unless he/she cannot find a leader in his/her neighborhood when he/she either makes his/her own decision or simply follows the crowd (details in **Following the leaders vs. following the crowd** in **Methods**).

The game with mixed leaders and followers is simulated in a small-world social network as given in Fig 2 with varying neighborhood sizes and percentages of leaders in the population. It is also assumed to be against a heterogeneous population as given in the example game in the previous section, where there are four population groups, and the first two groups contribute to the distancing risks differently from the last two. Table 4 contains some of the simulation results with the neighborhood size *k* = 3 but varying percentages of leaders in the population.

**Table 4.** Convergence of distancing strategies in mixed leader-follower populations.

| <1> % leaders | 10% | 20% | 30% | 40% | 50% | 60% |
| --- | --- | --- | --- | --- | --- | --- |
| $g_1: \langle \ x - x^*\rangle$ | 1.97e-02 | 1.70e-02 | 1.58e-02 | 1.09e-02 | 1.15e-02 | 8.36e-03 |
| $g_2: \langle \ x - x^*\rangle$ | 1.03e-02 | 1.17e-02 | 1.04e-02 | 7.48e-03 | 3.84e-03 | 2.42e-03 |
| $g_3: \langle \ x - x^*\rangle$ | 1.41e-02 | 1.26e-02 | 9.07e-03 | 8.10e-03 | 6.47e-03 | 2.62e-03 |
| $g_4: \langle \ x - x^*\rangle$ | 1.38e-02 | 1.29e-02 | 1.06e-02 | 7.98e-03 | 1.01e-03 | 5.87e-03 |
| <2> % leaders | 10% | 20% | 30% | 40% | 50% | 60% |
| $g_1: \langle \ x - x^*\rangle$ | 1.82e-03 | 1.40e-03 | 1.63e-03 | 1.96e-03 | 2.69e-03 | 4.14e-03 |
| $g_2: \langle \ x - x^*\rangle$ | 6.00e-04 | 7.40e-04 | 7.71e-04 | 1.03e-03 | 1.22e-03 | 1.27e-03 |
| $g_3: \langle \ x - x^*\rangle$ | 6.44e-04 | 6.15e-04 | 8.49e-04 | 9.68e-04 | 8.88e-04 | 8.03e-04 |
| $g_4: \langle \ x - x^*\rangle$ | 9.40e-04 | 1.09e-03 | 1.76e-03 | 3.37e-03 | 2.29e-03 | 2.78e-03 |

**Table Legends:** $x$ – individual strategy from simulation; $y$ – group strategy in neighborhood from simulation; $x^*$ – individual strategy at equilibrium; $y^*$ – group strategy at equilibrium; <1> – results by following the crowd if leaders not found; <2> – results by self-determination if leaders not found

The first set of results *<*1*>* in the table is obtained with a follow-the-crowd strategy if a follower cannot find a group leader among his/her closest neighbors. When there is a high percentage of leaders in the population (≥ 40%), the game converges well to the equilibrium strategies for all the individual groups. If there are lower than 20% leaders in the population, the convergence becomes less accurate (see top two plots in Fig 4), when most of the followers are not able to find a group leader in their neighborhood, and there is a disadvantage by simply following the crowd.

**Fig. 4.**
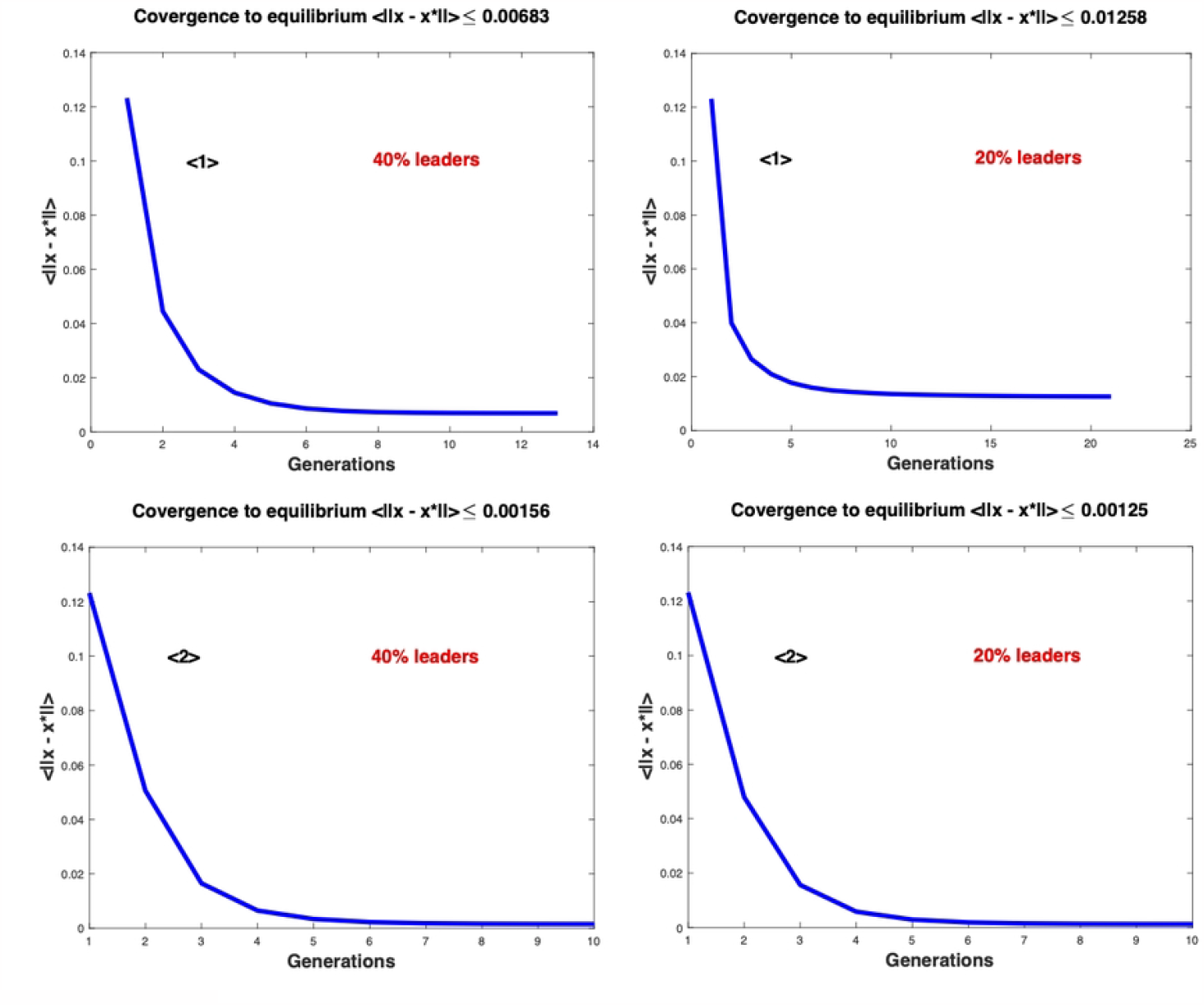
The convergence results for the distancing games with leaders. The difference between individual strategy *x* and equilibrium strategy *x*^∗^ is computed and averaged over all the individuals, and plotted at each generation. The top two graphs from left to right are the plots for the game with 40% and 20% of the individuals being assigned as leaders, and a follow-the-crowd strategy is used if a follower cannot find a group leader among his/her closest neighbors. The bottom two graphs from left to right are the plots for the game with 40% and 20% of individuals being assigned as leaders, and a make-own-decision strategy is used if a follower cannot find a group leader among his/her closest neighbors.

The second set of results *<*2*>* is obtained with a make-own-decision strategy if a follower cannot find a group leader among his/her closest neighbors. Similar to the previous case, when there is a high percentage of leaders in the population (≥ 40%), the game converges well to the equilibrium strategies for all the individual groups. In contrast to the previous case, when the percentage decreases to ≤20%, the convergence remains to be as accurate (see bottom two plots in Fig 4), because when there are fewer leaders, more followers start making their own decisions, which can be even better than following the leaders.

The simulation results with other neighborhood sizes and percentages of leaders (documented in Simulation results 3 in Supplementary Information) are all consistent with those in the above example. In general, when the percentage of leaders in the population is high, the game is expected to perform well and reach its equilibrium strategy. When the percentage is small, the game still runs reasonably well, showing that a small group of leaders are able to guide, and not every individual is required to make a decision. However, as shown in contrast between the set *<*1*>* and set *<*2*>* results, when there are fewer leaders, making own decisions actively is certainly more reliable than simply following the crowd.

## Discussion

The social distancing activities are not easy to track and hence difficult to study experimentally. The model proposed in this work presents a theoretical framework with which the distancing behaviors can be simulated, predicted, and analyzed. In this model, every individual in a given population is assumed to engage in social distancing by playing a distancing game, where based on what everybody else does, every individual makes decision on how to participate in a given set of social activities so that he/she can minimize his/her close social contacts with least possible negative social/economic impacts.

A distancing decision can be made to focus more on reducing close social contacts (*δ*_*i*_ *>* 0.5) or minimizing negative social/economic impacts (*δ*_*i*_ *<* 0.5). It depends on the severity of the epidemic and how the population responds to it. In general, if the epidemic is in a severe stage, a relatively large *δ*_*i*_ value would be good to reduce close social contacts, although the negative social/economic impacts may be high. If instead, a relatively small *δ*_*i*_ value is adopted, the amount of negative social/economic impacts is controlled, but the population would be exposed to more close social contacts. For example, for the distancing game demonstrated in Fig 1 in **Results** – **As a population game**, when in a severe epidemic, if *δ*_*i*_ is set to 1 for all *i*, the amount of close contacts for every individual is reduced to about 14 per week at equilibrium with the lost amount of necessary contacts around 35.21. However, if *δ*_*i*_ is set to 0 for all *i* instead, the amount of close contacts for every individual is increased to 59.95 per week, although the lost amount of necessary contacts is minimized to 14 per week at equilibrium.

The general distancing game assumes that every individual interacts with all others in the population. To be realistic, in this work, the population is assumed to be distributed over a social network, and each individual interacts only with his/her social neighbors. An algorithm is implemented to simulate the distancing game on such a network as discussed in **Results** – **In small-world social networks**. The algorithm is not equivalent to the general distancing game, and is not guaranteed to converge to an equilibrium strategy, either. In addition, the distancing game on a social network is played by every individual against his/her neighborhood, not the whole population. Nonetheless, the games simulated on social networks all converge to the equilibrium strategies of the general distancing games even when the neighborhood sizes are small and are restricted to contain only close neighbors, which is surprising, or not, as it may be how it is played out in the real world.

The extension of the distancing game to populations with multiple population groups in **Results** – **In heterogeneous populations** allows each population group to have its own assessment on distancing risks and hence its own distancing strategy. The equilibrium strategies of such games are justified in theory and also by simulation. In these games, the individuals take into account the contributions to their distancing risks from all population groups while giving more weights to those from their own groups, as they are supposed to interact more with the individuals in their own groups than those in other groups. However, the individuals may have more interactions with the individuals in some other groups as well as shown in several recent publications [98, 99], suggesting that in general, heavier weights may be assigned across more than one groups based on possible group-group interaction patterns.

The leadership role in social distancing is addressed lightly in this work as discussed in **Results** – **By following the leaders**. In fact, the leadership in social distancing is way beyond a matter of a few leaders making their own distancing decisions. For better or worse, leadership is often a determining factor in directing or changing the social distancing activities, as local or global organizations or governments make public health policies, provide social distancing guidelines, or give lockdown orders, etc., which are out of the scope of this study. However, the work in this study may help to understand the nature of social distancing as a collective behavior of human population, thereby providing a quantitative approach to assessing and improving the outcomes of public health policies concerning the control of social activities and its potential impacts.

The games discussed in this work are assumed to have only a small number of very general social activities, i.e., 20 CASA activities. In practice, there can be many more activities. They can also be of more specific types. For example, there can be different workplaces, different restaurants, and different shopping centers, and going to each of these places may be considered as a different social activity. The activities may also have some connections, i.e., not necessarily be independent of each other. An individual who participating in one of the activities may have contacts with individuals in other activities when the activities are carried out in close proximity in time or space. However, the proposed model can be extended to dependent activities by including the dependency among the activities in the estimation of the distancing risks [86].

The number of population groups in a heterogeneous population is not limited either, although only up to four population groups are considered in this work. The types of groups can also be combined. For example, groups can be formed according to the age as well as the vulnerability to the disease such as the vaccinated/unvaccinated children, recovered/unprotected adults, etc. In any case, the distancing activities in such populations can all be modeled as a multi-player game with each group corresponding to a single player and having its own risk assessment. In addition, in this work, the parameter *δ*_*i*_ is always set to a given value for all *i*, but in practice, it can be given a different value for a different *i*. For example, in certain situations, close contacts in schools or workplaces are less of a concern, and *δ*_*i*_ for these activities may therefore be given a smaller value than it is supposed to be. Further investigation into the use of this variation in practice can be pursued in future efforts.

## Methods

### Deriving contact and impact factors

The contact factor *α*_*i*_ in function *p*_*i*_ is the number of possible close contacts per unit time one may have in activity *i* when the activity is fully attended. It is not easy to estimate in general, for it may change from place to place or from time to time for a specific activity. In some sense, it is even unclear what a close contact means and how to measure it. In this work, only a general set of 20 CASA activities is considered. Their contact factors are determined by solving a so-called inverse game or inverse problem as demonstrated below.

First, the 20 CASA activities are grouped to 5 levels according their possible contact rates: The lowest level – activities 1-4; the second level – activities 5-8; the third level – activities 9-12; the fourth level – activities 13-16; and the fifth level – activities 17-20, according to common practices and CDC recommendations [26]. For each level of activities, a contact factor is to be assigned. Then, note that when *δ*_*i*_ = 1 for all *i*, the distancing game is equivalent to one for complete social distancing. Assume that the equilibrium strategy *x*^∗^ and hence *y*^∗^ in such a situation can be estimated. For example, with complete social distancing, an individual may stay home almost the whole day every day; he/she may spend 2 hours per day (or equivalently, 14 hours per week) for each of the first level activities, 1 hour per day (or equivalently, 7 hours per week) for each of the second, and so on and so forth (as listed in Table 5). Then, the contact factor *α*_*i*_ can be determined based on the condition *β*_*i*_*y*^∗^ = *λ* for some constant *λ* for all *i*. Set *α*_*i*_ to 1 for the first level activities to obtain *λ* = 1*/*8 contacts per hour (or equivalently, 14 contacts per week). The values for the rest of the *β*_*i*_ factors can then be extracted as given in Table 1.

**Table 5.** Estimated active times in 20 CASA activities for complete social distancing.

|  |  |  |  |  |  |  |  |  |  |  |
| --- | --- | --- | --- | --- | --- | --- | --- | --- | --- | --- |
| Act: | 1 | 2 | 3 | 4 | 5 | 6 | 7 | 8 | 9 | 10 |
| Time: | 14 | 14 | 14 | 14 | 7 | 7 | 7 | 7 | 4 | 4 |
| Act: | 11 | 12 | 13 | 14 | 15 | 16 | 17 | 18 | 19 | 20 |
| Time: | 4 | 4 | 2 | 2 | 2 | 2 | 1 | 1 | 1 | 1 |

Similarly, when *δ*_*i*_ = 0 for all *i*, the distancing game is equivalent to one where the individuals are free to choose their social activities as if there is no epidemics and no social distancing. Assume that the equilibrium strategy *x*^∗^ and hence *y*^∗^ in such a situation can be estimated. For example, in a week, if without social distancing, in average, an individual would work at home for 5 hours, stay with family for 5 hours, visit friends for 4 hours, and go to workplaces for 20 hours, etc. (as listed in Table 6). Then, the impact factor *β*_*i*_ can be determined based on the condition 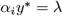 for some constant *λ* for all *i*. Choose *λ* to be the same level as for complete social distancing, i.e., *λ* = 1*/*8 contacts per hour (or equivalently, 14 contacts per week). Then, The values for all *β*_*i*_ can be extracted as given in Table 1.

**Table 6.** Estimated active times in 20 CASA activities when free of epidemics.

|  |  |  |  |  |  |  |  |  |  |  |
| --- | --- | --- | --- | --- | --- | --- | --- | --- | --- | --- |
| Act: | 1 | 2 | 3 | 4 | 5 | 6 | 7 | 8 | 9 | 10 |
| Time: | 5 | 5 | 3 | 3 | 5 | 3 | 3 | 5 | 5 | 3 |
| Act: | 11 | 12 | 13 | 14 | 15 | 16 | 17 | 18 | 19 | 20 |
| Time: | 3 | 5 | 4 | 4 | 16 | 20 | 6 | 6 | 4 | 4 |

Note that the active times assumed for the 20 CASA activities in Table 5 and Table 6 are based on common distancing practices. They are used to estimate the values for *α*_*i*_ and *β*_*i*_, which then define a plausible test case required for the analysis and simulation in this work. In real applications, they certainly need to be further refined and justified with more experimental data. Note also that the unit for *β*_*i*_ is the same as *α*_*i*_, i.e., the number of contacts per hour, but for *α*_*i*_ it is the number of close contacts one may have when fully participating in activity *i*, while for *β*_*i*_ it is the number of necessary contacts one may lose when staying only in this activity.

### Equilibrium strategies and stabilities

For each activity *i, p*_*i*_(*y*) = *w*_*i*_*y*_*i*_ with *w*_*i*_ = *δ*_*i*_*α*_*i*_ + (1− *δ*_*i*_)*β*_*i*_ and 0 ≤*δ*_*i*_ ≤1. If *w*_*i*_ *>* 0 for all *i*, the game reaches equilibrium when the potential distancing risks at all the activities are the same, i.e., an optimal strategy *x*^∗^ and hence *y*^∗^ is found such that 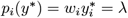 for some constant *λ* for all *i*. It follows that 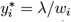 with 1*/λ* = Σ_*i*_ 1*/w*_*i*_ (proofs in **Appendix A1: General distancing games**). Note that 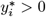 for all *i*, implying that the number of active activities is kept to the maximum (= *n*) at equilibrium.

Note also that the equilibrium strategy *x*^∗^ and hence *y*^∗^ is evolutionarily stable – a term used in evolutionary game theory [59, 60]. It means that if there is a small change in the strategy, the equilibrium strategy still prevails. In other words, if the population strategy *y*^∗^ is perturbed (or invaded) slightly by a new strategy *y, y*^∗^ will remain to be a better choice than *y*, and not be taken over by *y* (proofs in **Appendix A2: Evolutionary stability**).

### Generating small-world social networks

The social network is generated with the well-known Watts-Strogatz algorithm [89]. The algorithm has three parameters to determine a social network, *m* the number of the nodes, *K* the average degree of the nodes, and *b* the randomness of the connections. The algorithm generates a small-world social network of *m* nodes for a population of *m* individuals. In a cyclic order, the algorithm first connects each node with *K/*2 nodes next to the node on the right and then on the left. Then, for each node *i* and node *j* of *K/*2 nodes connected to node *i* on the right, the algorithm selects a node *k* not connected to *i*, and with a probability *b*, removes the link between *i* and *j* and connects *i* and *k*. In this way, the average degree of the nodes in the network would be around *K*, and the randomness of the connection between the connected nodes can be specified by *b*. A detailed algorithmic description for generating a small-world social network is given in Algorithm 1. A Matlab code can be found in the provided Simulation code 1, 2, 3 in Supplementary Information.

#### Algorithm 1

Generate a small-world network: (*m, K, b*)

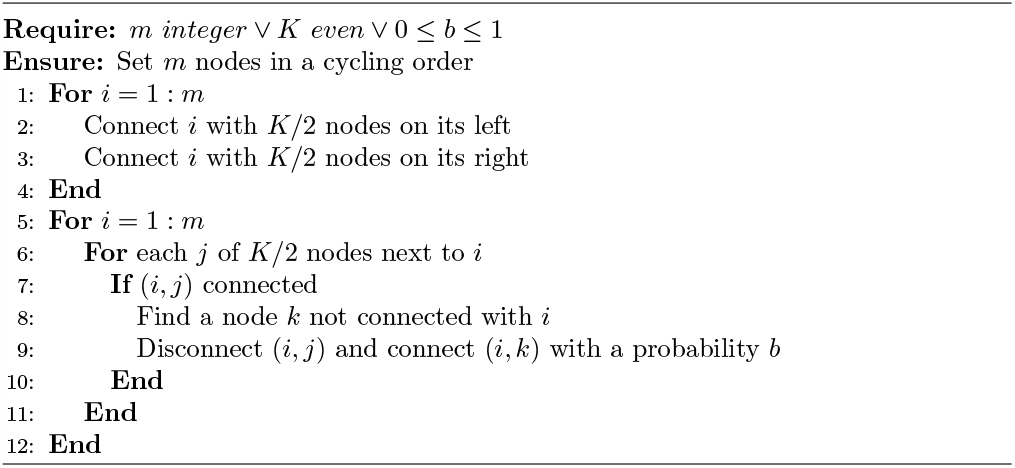

### Simulation of distancing games in social networks

The simulation of distancing games is based on the general principle of replicator dynamics for population games [59, 60]. If *y*_*i*_ is the participating frequency of the population in activity *i* at a certain time *t*, the replicator dynamics states that the changing rate of *y*_*i*_ is proportional to the difference between the potential distancing risk *p*_*i*_(*y*) at activity *i* and the population average Σ_*i*_ *y*_*i*_*p*_*i*_(*y*). If the potential distancing risk is higher than the average, activity *i* is considered to be riskier, and *y*_*i*_ (and hence *x*_*i*_) should be decreased; otherwise, *y*_*i*_ (and hence *x*_*i*_) should be increased.

For the game on a social network, the simulation can be done for every individual against the population in his/her neighborhood. The population strategy *y* then becomes the neighborhood strategy, which is different for a different individual in general. The neighborhood of size *k* of an individual includes all the neighbors connected to the individual with up to *k* consecutive connections. Depending on the neighborhood size, the game becomes against a fraction of population ranging from the immediate neighbors of the individual to the whole population. Algorithm 2 gives more algorithmic description on how an individual updates his/her strategy in every round of the game. A Matlab code for the whole simulation is provided (in Simulation code 1 in Supplementary Information).

The simulation starts with a strategy for every individual randomly generated around its equilibrium one. More specifically, if 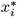 is the equilibrium value of the participating frequency for activity *i*, then first perturb 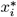 randomly by a certain percentage, say 20%, and then generate *x*_*i*_ randomly within 80% of deviation from the perturbed value of 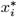. The simulation proceeds in multiple generations. At each generation, every individual plays the game once, i.e., has a chance to update his/her strategy. The simulation ends when either every individual strategy converges to the equilibrium strategy in average or it stops making any further progress. Every simulation is repeated for 5 times and an average output is recorded and reported.

### Distancing in heterogeneous populations

Consider a simple case where the population is divided into two groups, groups *a* and *b*. Let *x*^*a*^ and *x*^*b*^ be the distancing strategies for individuals in groups *a* and *b*, respectively, with 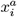 and 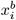 being the participating frequencies of the individuals in activity *i*. Let *y*^*a*^ and *y*^*b*^ be the average strategies of the individuals in groups *a* and *b* in the population, with 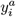and 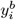 being the corresponding average participating frequencies of these individuals in activity *i*. Given strategies *y*^*a*^ and *y*^*b*^ in the population, the potential distancing risk at activity *i* can be estimated by a function 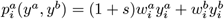 for a group *a* individual or 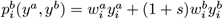 for a group *b* individual, where *w*^*a*^ and *w*^*b*^ are risk factors for groups *a* and *b*, respectively, and the contribution to the distancing risk from the individual’s own group is given more weight (1 + *s*) for some *s >* 0, as the individual is supposed to interact more with the individuals in his/her own group than those in the other group.

Then, for an individual of strategy *x*^*a*^ in group *a*, the distancing risk to participate in given *n* activities can be evaluated by a function 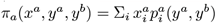. Similarly, for an individual of strategy *x*^*b*^ in group *b*, the distancing risk to participate in given *n* activities can be evaluated by a function 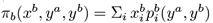). Together, with these functions, a multi-player distancing game can be defined for the whole population with each population group corresponding to a single player; and a pair of strategies *x*^*a*∗^ and *x*^*b*∗^ form an equilibrium pair of strategies for the game if and only if *y*^*a*∗^ = *x*^*a*∗^ and *y*^*b*∗^ = *x*^*b*∗^, and for every individual in group *a*, the distancing risk *π*_*a*_(*x*^*a*∗^, *y*^*a*∗^, *y*^*b*∗^) using strategy *x*^*a*∗^ is no greater than the distancing risk *π*_*a*_(*x*^*a*^, *y*^*a*∗^, *y*^*b*∗^) using any other strategy *x*^*a*^, and for every individual in group *b*, the distancing risk *π*_*b*_(*x*^*b*∗^, *y*^*a*∗^, *y*^*b*∗^) using strategy *x*^*b*∗^ is no greater than the distancing risk *π*_*b*_(*x*^*b*^, *y*^*a*∗^, *y*^*b*∗^) using any other strategy *x*^*b*^. It follows from a little bit analysis that the equilibrium strategy for each

#### Algorithm 2

Updating individual distancing strategies

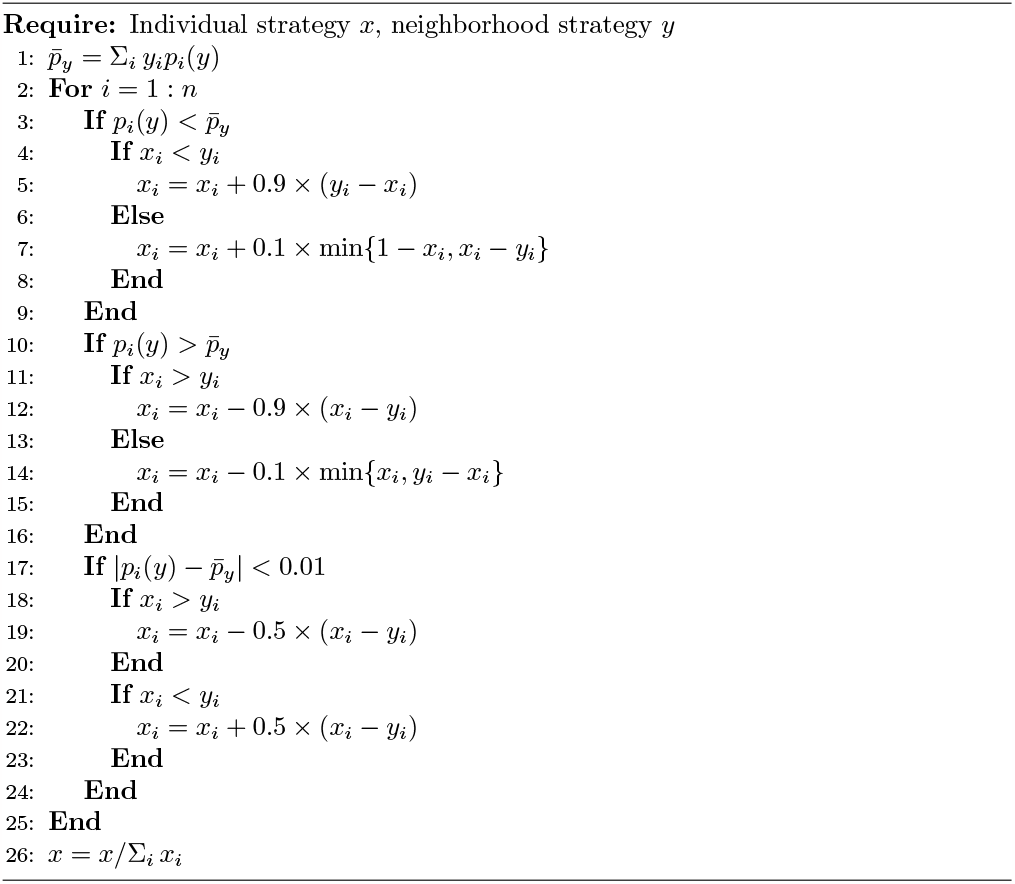

population group can be obtained with 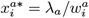 for all *i* for some constant *λ*_*a*_ such that 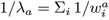, and 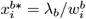 for all *i* for some constant *λ*_*b*_ such that 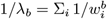. At equilibrium, *y*^*a*∗^ = *x*^*a*∗^ and *y*^*b*∗^ = *x*^*b*∗^, and the average population strategy should be *y*^∗^ = *ρ*_*a*_*y*^*a*∗^ + *ρ*_*b*_*y*^*b*∗^, where *ρ*_*a*_ and *ρ*_*b*_ are the percentages of group *a* and *b* individuals in the population, respectively. These results can be extended straightforwardly to populations with more than two population groups (general descriptions and proofs in **Appendix A3: Games with multiple population groups**).

The simulation of the distancing game with multiple population groups is done with the population also distributed over a small-world social network, where the game is played by every individual against his/her population group in his/her neighborhood. The key difference of this simulation from the one described in Algorithm 2 is that the potential distancing risk at each activity is estimated using a formula as described above, and the average risk over all activities is evaluated for each individual using his/her group strategy in his/her neighborhood. A Matlab code for simulating the distancing games with up to four population groups is provided (in Simulation code 2 in Supplementary Information).

### Following the leaders vs. following the crowd

A certain percentage of individuals are randomly selected as leaders. A leader makes distancing decisions as a regular player for the distancing game, whether the game is played in a small-world social network or with multiple population groups. A follower tries to find the leaders in his/her population group in his/her neighborhood, and copies the average strategy of the leaders. If he/she fails to find a leader among his/her closest neighbors, he/she either makes her own decision as a regular player or follows the crowd by copying the average strategy of his/her group members in his/her neighborhood. The simulation is done with the percentage of leaders in the population varying from 10% to 20%, 30%, 40%, 50%, and 60%, and the neighborhood size changing from 1 to 2, 3, 4, 5, and 6. The parameters for the network are fixed to *m* = 200, *K* = 6, and *b* = 0.3. A Matlab code for the simulation is provided (in Simulation code 3 in Supplementary Information).

## Supplementary information

Results from computer simulation conducted in this work are documented in the following files. The Matlab codes producing the results are all provided.

Simulation results 1: SI-simulation-results-1.pdf

Simulation results 2: SI-simulation-results-2.pdf

Simulation results 3: SI-simulation-results-3.pdf

Simulation code 1: run_simulation 1.m; simulation 1.m

Simulation code 2: run_simulation 2.m; simulation 2.m

Simulation code 3: run_simulation 3.m; simulation 3.m

## Data Availability

All relevant data are within the manuscript and its Supporting Information files.

## Acknowledgments

The author would like to acknowledge the support from the Simons Foundation on this work through the Mathematics and Physical Sciences Collaboration Grants for Mathematicians (Award Number: 586065).

## Appendix A

### Appendix A1: General distancing games

Assume that the population has *n* activities. Let *x* = {*x*_*i*_ : *i* = 1, …, *n*} be a set of frequencies representing the distancing strategy of any individual, with *x*_*i*_ being the frequency of the individual to participate in activity *i*, and Σ_*i*_ *x*_*i*_ = 1. Let *y* = {*y*_*i*_ : *i* = 1, …, *n*} be a set of frequencies representing the strategy of the population, with *y*_*i*_ being the average frequency of all the individuals in the population to participate in activity *i*, and Σ_*i*_ *y*_*i*_ = 1.

Given a distancing strategy *y* from the population, assume that each individual can estimate the potential distancing risk at each activity *i* using a function *p*_*i*_(*y*). Then, the distancing risk of the individual of strategy *x* at activity *i* must be *x*_*i*_*p*_*i*_(*y*), and at all the activities together be Σ_*i*_ *x*_*i*_*p*_*i*_(*y*) = *π*(*x, y*).

#### Definition 1 (Distancing game).

*A distancing game is a population game where every individual chooses a strategy x against a strategy y of the population so that his/her distancing risk π*(*x, y*) *can be minimized*.

#### Definition 2 (Equilibrium strategy).

*A strategy x*^∗^ *is an equilibrium strategy of the distancing game if and only if every individual in the population takes this strategy x*^∗^ *(and hence y*^∗^ = *x*^∗^*) such that his/her distancing risk π*(*x*^∗^, *y*^∗^) ≤*π*(*x, y*^∗^) *for any strategy x*.

#### Theorem 1.

*A strategy x*^∗^ *is an equilibrium strategy for the distancing game if and only if there is a constant λ such that*

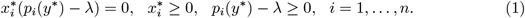

*Proof*. (=*>*) Suppose that *x*^∗^ is an equilibrium strategy. Then, *π*(*x*^∗^, *y*^∗^) ≤ *π*(*x, y*^∗^) for any strategy *x*, and therefore, *π*(*x*^∗^, *y*^∗^) ≤ *π*(*e*_*i*_, *y*^∗^) = *p*_*i*_(*y*^∗^), *i* = 1, …, *n*, where *e*_*i*_ is the *i*th unit vector. Let *π*(*x*^∗^, *y*^∗^) = *λ*. Then *p*_*i*_(*y*^∗^) − *λ* ≥ 0 for all *i* = 1, …, *n*. For any *i*, if 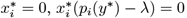; if 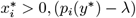 must be zero, for otherwise, 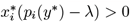. Collect the latter inequality for all *i* to obtain 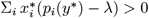. Then *π*(*x*^∗^, *y*^∗^) −*λ >* 0, which is contradictory to the fact that *π*(*x*^∗^, *y*^∗^) = *λ*. Thus the conditions in (1) are all satisfied.

(*<*=) Suppose there is a parameter *λ* such that *x*^∗^ satisfies all the conditions in (1).

Collect the first equation in (1) for all *i* to obtain 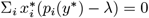, which is equivalent to *π*(*x*^∗^, *y*^∗^) −*λ* = 0, and therefore, *λ* = *π*(*x*^∗^, *y*^∗^). Let *x* be an arbitrary strategy. Multiply the last equation in (1) by *x*_*i*_ to obtain *x*_*i*_(*p*_*i*_(*y*^∗^)− *λ*) ≥ 0. Collect the latter inequality to obtain Σ_*i*_ *x*_*i*_(*p*_*i*_(*y*^∗^) −*λ*) ≥ 0, which is equivalent to *π*(*x, y*^∗^) −*λ* ≥ 0. Then, *π*(*x*^∗^, *y*^∗^) ≤*π*(*x, y*^∗^) for any strategy *x*, and *x*^∗^ is an equilibrium strategy.

#### Theorem 2.

*Assume that the activities are independent and function p*_*i*_(*y*) = *w*_*i*_*y*_*i*_ *with w*_*i*_ *>* 0 *for all i. Then*, 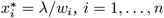, *form an equilibrium strategy for the distancing game, where λ is a constant such that* 1*/λ* = Σ_*i*_ 1*/w*_*i*_.

*Proof*. Let 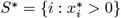. Then, by Theorem 1, for *i* ∈ *S*^∗^, *p*_*i*_(*y*^∗^) − *λ* = 0 for some constant *λ*. It follows that 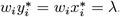, and 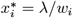 for all *i* ∈ *S*^∗^. Since the sum of all 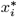 equals 1, the sum of the latter equations gives 1*/λ* = Σ_*i*∈*S*∗_ 1*/w*_*i*_. Note that *S*^∗^ must contain all *i* = 1, …, *n*, for otherwise, if there is *i* such that 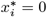, then 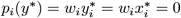; it follows that *p*_*i*_(*y*^∗^) *< λ*, which contradicts to the fact that *p*_*i*_(*y*^∗^) ≥ *λ* for all *i* by Theorem 1.

### Appendix A2: Evolutionary stability

#### Definition 3

(Evolutionary stability). *An equilibrium strategy x*^∗^ *for the distancing game is evolutionarily stable if for any strategy x* ≠ *x*^∗^, *there is* 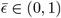 *such that π*(*x*^∗^, *Ex* + (1 − *E*)*x*^∗^) *< π*(*x, Ex* + (1 − *E*)*x*^∗^) *for all* 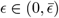 *[59, 60]*.

#### Definition 4 (Potential minimization).

*Let f* (*y*) *be a function such that* 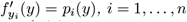. *Then, the problem*

min{*f* (*y*) : Σ_*i*_ *y*_*i*_ = 1, *y*_*i*_ ≥ 0, *i* = 1, …, *n*} *is called a potential minimization problem for the distancing game defined by p*_*i*_(*y*), *i* = 1, …, *n [100, 101]*.

#### Theorem 3.

*A strategy x*^∗^ *is an equilibrium strategy for the distancing game if and only if x*^∗^ *is a KKT point of the corresponding potential minimization problem*.

#### Theorem 4.

*An equilibrium strategy x*^∗^ *for the distancing game is evolutionarily stable if and only if x*^∗^ *is a strict local minimizer of the corresponding potential minimization problem*.

#### Theorem 5.

*Assume that the activities are independent and function p*_*i*_(*y*) = *w*_*i*_*y*_*i*_ *with w*_*i*_ *>* 0 *for all i. Then, the equilibrium strategy* 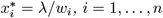, *for the distancing game is evolutionarily stable with* 1*/λ* = Σ_*i*_ 1*/w*_*i*_.

*Proof*. The Hessian of the objective function *f* (*x*) of the potential minimization problem corresponding to the distancing game is a diagonal matrix with *w*_*i*_, *i* = 1, …, *n* as the diagonal elements. Since *w*_*i*_ *>* 0 for all *i*, the Hessian is positive definite, which guarantees the solution to the potential minimization problem *x*^∗^ to be a strict local minimizer. It follows that *x*^∗^ must be evolutionarily stable by Theorem 4.

### Appendix A3: Games with multiple population groups

Assume that the population is divided into *M* groups. Let *x*^(*j*)^ be the strategy of an individual in group *j*, and *y*^(*j*)^ the average strategy of all group *j* individuals in the population. Let 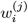 be the risk factor for activity *i* for the individuals in group *j*. Then, the potential distancing risk for a group *j* individual at activity *i* can be defined as

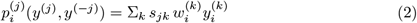

where *y*^(−*j*)^ represents all group strategies *y*^(1)^, …, *y*^(*M*)^ excluding *y*^(*j*)^, Σ_*k*_ means the sum over all *k* = 1, …, *M*, and *s*_*jk*_ is a scaling factor, *s*_*jk*_ = (1 + *s*) for some *s >* 0 if *k* = *j* and *s*_*jk*_ = 1 if *k≠j*, thus the contribution of group *j* to the distancing risk is given more weight as an individual in group *j* is supposed to interact more with the individuals in his/her own group than those in other groups.

#### Definition 5

(Distancing game with multiple population groups). *Assume that the population is divided into M groups. Let* 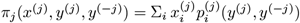. *j* = 1, … *M* Then together, with all these functions, a multi-player distancing game can be formed with each population group corresponding to a single player; and a set of strategies *x*^(*j*)∗^, *j* = 1, …, *M, is an equilibrium set of strategies for the game if and only if for all j* = 1, …, *M, y*^(*j*)∗^ = *x*^(*j*)∗^, *and π*_*j*_(*x*^(*j*)∗^, *y*^(*j*)∗^, *y*^(−*j*)∗^) ≤ *π*_*j*_(*x*^(*j*)^, *y*^(*j*)∗^, *y*^(−*j*)∗^) *for any strategy x*^(*j*)^.

#### Theorem 6.

*Assume that the population is divided into M groups and the activities are independent. Assume that the function for an individual in group j to evaluate the potential distancing risk at activity i is given by (2) with* 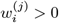 *for all i and j*.

*Then, there is a unique set of equilibrium strategies x*^(*j*)∗^, *j* = 1, …, *M, for the multi-player distancing game of the population*, 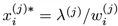 *for all i, where λ*^(*j*)^ *is a constant such that* 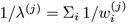.

*Proof*. Let *x*^(*j*)∗^ be the strategy of an individual in group *j* at equilibrium and *y*^(*j*)∗^ the average strategy of group *j* individuals in the population. Then, it is necessary and sufficient that for each group *j*, 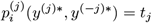, *y*^(−*j*)∗^) = *t*_*j*_ for all *i* for some constant *t*_*j*_, i.e.,

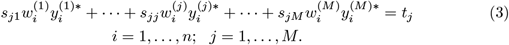

The above equations can be written in a more compact form as:

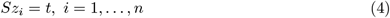

where *S* = (*s*_*jk*_) is an *M* ×*M* matrix, *s*_*jk*_ = (1 + *s*) for some *s >* 0 if *j* = *k* and *s*_*jk*_ = 1 if *j* ≠ *k, z*_*i*_ and *t* are *M* -dimensional vectors, 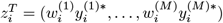, and *t*^*T*^ = (*t*_1_, …, *t*_*M*_). It is not difficult to verify that *S* is nonsingular. Therefore, *z*_*i*_ = *S*^−1^*t*. Let (*S*^−1^*t*)_*j*_ = *λ*^(*j*)^, *j* = 1, …, *M*. Then, 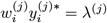 for all *i*. It follows that 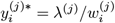 with 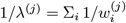.

## References

1. R. J. Hatchett, C. E. Mecher, and M. Lipsitch, Public health interventions and epidemic intensity during the 1918 influenza pandemic, PNAS 104: 7582–7587, 2007, www.pnas.org/cgi/doi/10.1073/pnas.0610941104

2. P. Caley, D. J. Philp, and K. McCracken, Quantifying social distancing arising from pandemic influenza, Journal of the Royal Society Interface 5: 631–639, 2008, 10.1098/rsif.2007.1197

3. D. Roth and B. Henry, Social distancing as a pandemic influenza prevention measure, National Collaborating Center for Infectious Diseases, Winnipeg, MB, Canada, 2011, http://www.nccid.ca

4. F. Ahmed, N. Zviedrite, and A. Uzicanin, Effectiveness of workplace social distancing measures in reducing influenza transmission: A systematic review, BMC Public Heath 8: 518–530, 2018, 10.1186/s12889-018-5446-1

5. O. B. Cano, S. C. Morales, and C. Bendtsen, COVID-19 modeling: The effects of social distancing, Interdisciplinary Perspectives on Infectious Diseases, 2020, Article ID 2041743, 10.1155/2020/2041743

6. N. M. Ferguson, D. Laydon, G. Nedjati-Gilani, et al., Impact of non-pharmaceutical interventions (NPIs) to reduce COVID-19 mortality and healthcare demand, Imperial College London, 2020, 10.25561/77482

7. M. Greenstone and V. Nigam, Does social distancing matter? Covid Economics 7: 1-22, 2020

8. N. Islam, S. J. Sharp, G. Chowell, et al., Physical distancing interventions and incidence of coronavirus disease 2019: natural experiment in 149 countries, BMJ 370: m2743, 2020, 10.1136/bmj.m2743

9. N. B. Masters, S. F. Shih, A. Bukoff, et al., Social distancing in response to the novel coronavirus (COVID-19) in the United States, PLoS ONE 15: e0239025, 2020, 10.1371/journal.pone.0239025

10. D. J. McGrail, J. Dai, K. M. McAndrews, and R Kalluri, Enacting national social distancing policies corresponds with dramatic reduction in COVID19 infection rates, PLoS ONE 15(7): e0236619, 2020, 10.1371/journal.pone.0236619

11. M. J. Siedner, G. Harling, Z. Reynolds, et al., Social distancing to slow the US COVID-19 epidemic: Longitudinal pretest-posttest comparison group study, PLoS Medicine 17: e1003244, 2020, 10.1371/journal.pmed.1003244

12. A. Nande, B. Adlam, J. Sheen, et al., Dynamics of COVID-19 under social distancing measures are driven by transmission network structure, PLoS Computational Biology 17: e1008684, 2021, 10.1371/journal.pcbi.1008684

13. G. A. Wellenius, S. Vispute, V. Espinosa, et al., Impacts of social distancing policies on mobility and COVID-19 case growth in the US, Nature Communications 12: 3118, 2021, 10.1038/s41467-021-23404-5

14. S. Kissler, C. Tedijanto, E. Goldstein, et al., Projecting the transmission dynamics of SARS-CoV-2 through the post-pandemic period, Science 368: 860–868, 2020, 10.1126/science.abb5793

15. C. Mann, Pandemics leave us forever altered – What history can tell us about the long-term effects of the coronavirus, The Atlantic: IDEAS, June 2020, https://www.theatlantic.com/magazine/archive/2020/06/pandemics-plagues-history/610558

16. A. B. Abel and S. Panageas, Social distancing, vaccination and the paradoxical optimality of an endemic equilibrium, NBER Working Paper 27742, 2021, http://www.nber.org/papers/w27742

17. L. Dyson, E. M. Hill, S. Moore, et al., Possible future waves of SARS-CoV-2 infection generated by variants of concern with a range of characteristics, Nature Communications 12: 5730, 2021, 10.1038/s41467-021-25915-7

18. M. Galanti, S. Pei, T. K. Yamana, et al., Social distancing remains key during vaccinations, Science 371: 473–474, 2021, 10.1126/science.abg4380

19. Z. M. Afshar, M. Barary, R. Hosseinzadeh, et al., Breakthrough SARS-CoV-2 infections after vaccination: a critical review, Human Vaccines & Immunotherapeutics 18: e2051412, 2022, 10.1080/21645515.2022.2051412

20. J. M. Barrero, N. Bloom, and S. J. Davis, Long social distancing, NBER Working Paper 30568, 2022, http://www.nber.org/papers/w30568

21. L. O. Gostin, Life after the COVID-19 pandemic, JAMA Health Forum 3: e220323, 2022, 10.1001/jamahealthforum.2022.0323

22. S. Saha, G. Samanta, and J. J. Nieto, Impact of optimal vaccination and social distancing on COVID-19 pandemic, Mathematics and Computers in Simulation 200: 285–314, 2022, 10.1016/j.matcom.2022.04.025

23. N. R. Jones, Z. U. Qureshi, R. J. Temple, et al., Two meters or one: what is the evidence for physical distancing in covid-19? BMJ 370: m3223, 2020, 10.1136/bmj.m3223

24. K. Pearce, What is social distancing and how can it slow the spread of COVID-19?, HUB, Johns Hopkins University, 2020, https://hub.jhu.edu/2020/03/13/what-is-social-distancing

25. M. Z. Bazant and J. W. M. Bush, A guideline to limit indoor airborne transmission of COVID-19, PNAS 118: e2018995118, 2021, 10.1073/pnas.2018995118

26. CDC US, Isolation and precautions for people with COVID-19, CDC COVID-19 Guidelines, 2022, https://www.cdc.gov/coronavirus/2019-ncov/your-health/isolation.html

27. R. Glass, L. Glass, W. Beyeler, and H. Min, Targeted social distancing design for pandemic influenza, Emerging Infectious Diseases 12: 1671–1681, 2006, http://www.cdc.gov/eid

28. J. Kelso, G. Milne, and H. Kelly, Simulation suggests that rapid activation of social distancing can arrest epidemic development due to a novel strain of influenza, BMC Public Health 9: 117, 2009, http://www.biomedcentral.com/1471-2458/9/117

29. Y. T. Yang and R. D. Silverman, Social distancing and the unvaccinated, The New England Journal of Medicine 372: 1481–1483, 2015, 10.1056/NEJMp1501198

30. L. Faherty, H. Schwartz, F. Ahmed, et al., School and preparedness officials’ perspectives on social distancing practices to reduce influenza transmission during a pandemic: Considerations to guide future work, Preventive Medicine Reports 14: 100871, 2019, 10.1016/j.pmedr.2019.100871

31. C. Adolph, K. Amano, B. Bang-Jensen, et al., Pandemic politics: Timing state-level social distancing responses to COVID-19, Center for Statistics and the Social Sciences, University of Washington, medRxiv preprint, 10.1101/2020.03.30.20046326

32. C. Benke, L. K. Autenrieth, E. Asselmann, and C. A. Pane-Farre, Lockdown, quarantine measures, and social distancing: Associations with depression, anxiety and distress at the beginning of the COVID-19 pandemic among adults from Germany, Psychiatry Research 293: 113462, 2020, 10.1016/j.psychres.2020.113462

33. P. Block, M. Hoffman, I. J. Raabe, Social network-based distancing strategies to flatten the COVID-19 curve in a post-lockdown world, Nature Human Behaviour 588: 588–596, 2020, 10.1038/s41562-020-0898-6

34. H. Carel, M. Ratcliffe, and T. Froese, Reflecting on experiences of social distancing, The Lancet 396: 87–88, 2020, 10.1016/S0140-6736(20)31485-9

35. L. Cerbara, G. Ciancimino, M. Crescimbene, et al., A nation-wide survey on emotional and psychological impacts of COVID-19 social distancing, European Review for Medical and Pharmacological Sciences 24: 7155–7163, 2020

36. S. Gupta, K. I. Simon, C. Wing, Mandated and voluntary social distancing during the COVID-19 epidemic: A review, NBER Working Paper 28139, 2020, http://www.nber.org/papers/w28139

37. A. Jawaid, Protecting older adults during social distancing, Science 368: 145, 2020, 10.1126/science.abb7885

38. K. A. Van Orden, E. Bower, J. Lutz, et al., Strategies to promote social connections among older adults during “social distancing” restrictions, The American Journal of Geriatric Psychiatry 29: 816–827, 2020, 10.1016/j.jagp.2020.05.004

39. A. Strong and J. W. Welburn, An estimation of the economic costs of social-distancing policies, Research Report, RAND Corporation, 2020, http://www.rand.org/t/rra173-1

40. A. Wilder-Smith and D. Freedman, Isolation, quarantine, social distancing and community containment: pivotal role for old-style public heath measures in the novel coronavirus (2019-nCoV) outbreak, Journal of Travel Medicine 2020: 1–4, 10.1093/jtm/taaa020

41. Z. Barnett-Howell, O. J. Watson, and A. M. Mobarak, The benefits and costs of social distancing in high- and low-income countries, Trans R Soc Trop Med 0: 1–13, 2021, 10.1093/trstmh/traa140

42. E. Y. Choi, M. P. Farina, Q. Wu, and J. Ailshire, COVID-19 social distancing measures and loneliness among older adults, Journal of Gerontology: Social Sciences 77: e167–e178, 2021, 10.1093/geronb/gbab009

43. M. Farboodi, G. Jarosch, and R. Shimer, Internal and external effects of social distancing in a pandemic, Journal of Economic Theory 196: 105293, 2021, 10.1016/j.jet.2021.105293

44. R. I. Mukhamadiarov, S. Deng, S. R. Serrao, et al., Requirements for the containment of COVID-19 disease outbreaks through periodic testing, isolation, and quarantine, Journal of Physics A: Mathematical and Theoretical 55: 034001, 2021, 10.1088/1751-8121/ac3fc3

45. R. Vardavas, P. N. de Lima, P. K. Davis, et al., Modeling infectious behaviors: The need to account for behavioral adaptation in COVID-19 models, Policy Complex Sys. 7: 21–32, 2021, 10.18278/jpcs.7.1.3

46. A. Aleman and I. Sommer, The silent danger of social distancing, Psychological Medicine 52: 789–790, 2022, 10.1017/S0033291720002597

47. D. Bzdok and R. I. M. Dunbar, Social isolation and the brain in the pandemic era, Nature Human Behaviour 6: 1333–1343, 2022, 10.1038/s41562-022-01453-0

48. O. Delardas, K. S. Kechagias, P. N. Pontikos, and P. Giannos, Socio-economic impacts and challenges of the coronavirus pandemic (COVID-19): An updated review, Sustainability 14: 9699, 2022, 10.3390/su14159699

49. T. Rossetti, S. Y. Yoon, and R. A. Daziano, Social distancing and store choice in times of a pandemic, Journal of Retailing and Consumer Services 65: 102860, 2022, 10.1016/j.jretconser.2021.102860

50. S. Sims, R. Harris, S. Hussein, et al., Social distancing and isolation strategies to prevent and control the transmission of COVID-19 and other infectious diseases in care homes for older people: An international review, International Journal of Environmental Research and Public Health 19: 3450, 2022, 10.3390/ijerph19063450

51. M. J. Tildesley, A. Vassall, S. Riley Steven, et al., Optimal health and economic impact of non-pharmaceutical intervention measures prior and post vaccination in England: A mathematical modeling study, Royal Society Open Science 9: 211746, 2022, 10.1098/rsos.211746

52. T. Abel and D. McQueen, The COVID-19 pandemic calls for spatial distancing and social closeness: not for social distancing! International Journal of Public Health 65: 231, 2020, 10.1007/s00038-020-01366-7

53. J. Lammers, J. Crusius, and A. Gast, Correcting misperceptions of exponential coronavirus growth increases support for social distancing, PNAS 117: 16264–16266, 2020, https://www.pnas.org/cgi/doi/10.1073/pnas.2006048117

54. N. J. Long, From social distancing to social containment, Medicine Anthropology Theory 7: 247–260, 2020, 10.17157/mat.7.2.791.

55. G. Miller, Social distancing prevents infections, but it can have unintended consequences, Science News, March 16, 2020, 10.1126/science.abb7506

56. C. J. Tyrrell and K. N. Williams, The paradox of social distancing: Implications for older adults in the context of COVID-19, Psychological Trauma: Theory, Research, Practice, and Policy, 2020, 10.1037/tra0000845

57. S. N. Williams, C. J. Armitage, T. Tampe, and K. Dienes, Public perceptions and experiences of social distancing and social isolation during the COVID-19 pandemic: a UK-based focus group study, BMJ Open 10: e039334, 2020, 10.1136/bmjopen-2020-039334

58. C. E. Kontokosta, B. Hong, and B. J. Bonczak, Measuring sensitivity to social distancing behavior during the COVID-19 pandemic, Nature Scientific Reports, 2022, 10.1038/s41598-022-20198-4

59. J. W. Weibull Evolutionary Game Theory, The MIT Press, 1995

60. J. Hofbauer and K. Sigmund, Evolutionary Games and Population Dynamics, Cambridge University Press, 1998

61. T. Reluga, Game theory of social distancing in response to an epidemic, PLoS Computational Biology 6: c1000793, 2010, 10.1371/journal.pcbi.1000793

62. L. Valdez, C. Buono, P. Macri, and L. Braunstein, Intermittent social distancing strategies for epidemic control, Physics Review E. 85: 036108, 2012, 10.1103/PhysRevE.85.036108

63. E. Fenichel, Economic considerations of social distancing and behavioral based policies during an epidemic, Journal of Health Economics 32: 440–451, 2013, 10.1016/j.jhealeco.2013.01.002

64. T. Reluga, Equilibria of an epidemic game with piecewise linear social distancing cost, Bulletin of Mathematical Biology 75: 1961–1984, 2013, 10.1007/s11538-013-9879-5

65. T. Britton, F. Ball, and P. Trapman, A mathematical model reveals the influence of population heterogeneity on herd immunity to SARS-CoV-2, Science 369: 846–849, 2020, 10.1126/science.abc6810

66. S. Cato, T. Iida, K. Ishida, et al., Social distancing as a public good under the COVID-19 pandemic, Public Health 88: 51–53, 2020, 10.1016/j.puhe.2020.08.005

67. A. Glaubitz and F. Fu, Oscillatory dynamics in the dilemma of social distancing, Proceedings of Royal Society A 476: 20200686, 2020, 10.1098/rspa.2020.0686

68. T. Kruse and P. Strack, Optimal control of an epidemic through social distancing, Cowles Foundation Discussion Paper Number 2229R, Yale University, 2020, https://papers.ssrn.com/sol3/papers.cfm?abstractid=3581295

69. C. J. E. Metcalf, D. H. Morris, and S. W. Park, Mathematical models to guide pandemic response, Science 369: 368–369, 2020, 10.1126/science.abd1668

70. C. M. Peak, R. Kahn, Y. H. Grad, L. M. Childs, et al., Modeling the comparative impact of individual quarantine vs. active monitoring of contacts for the mitigation of COVID-19, The Lancet Infectious Diseases 20: 1025–1033, 2020, 10.1016/S1473-3099(20)30361-3.

71. F. Toxvaerd, Equilibrium social distancing, Cambridge-INET Working Paper Series No. 2020/08, University of Cambridge, 2020

72. S. Bugalia, J. P. Tripathi, and H. Wang, Mathematical modeling of intervention and low medical resource availability with delays: applications to COVID-19 outbreaks in Spain and Italy, Mathematical Biosciences and Engineering 18: 5865–5920, 2021, 10.3934/mbe.2021295

73. E. M. Hill, B. D. Atkins, M. J. Keeling, L. Dyson, and M. J. Tildesley, A network modeling approach to assess non-pharmaceutical disease controls in a worker population: An application to SARS-CoV-2, PLoS Computational Biology 17: e1009058, 2021, 10.1371/journal.pcbi.1009058

74. X. Luo, S. Feng, J. Yang, et al., Non-pharmaceutical interventions contribute to the control of COVID-19 in China based on a pairwise model, Infectious Disease Modeling 6: 643–663. 2021, 10.1016/j.idm.2021.04.001

75. H. Amini and A. Minca, Epidemic spreading and equilibrium social distancing in heterogeneous networks, Dynamic Games and Applications 12: 258–287, 2022, 10.1007/s13235-021-00411-1

76. M. A. Ovi, K. N. Nabi, K. M. A. Kabir, Social distancing as a public-good dilemma for socio-economic cost: An evolutionary game approach, Heliyon 8: e11497, 2022, 10.1016/j.heliyon.2022.e11497

77. K. Chen, C. S. Pun, and H. Y. Wong, Efficient social distancing during the COVID-19 pandemic: Integrating economic and public health considerations, European Journal of Operational Research 304: 84–98, 2023, 10.1016/j.ejor.2021.11.012

78. S. A. Nowak, P. N. de Lima, and R. Vardavas, Optimal non-pharmaceutical pandemic response strategies depend critically on time horizons and costs, Scientific Reports 3: 2416, 2023, 10.1038/s41598-023-28936-y

79. J. Maynard-Smith and G. R. Price, The logic of animal conflict, Nature 246: 15–18, 1973

80. R. Axelrod and W. D. Hamilton, The evolution of cooperation, Science 211: 1390–1396, 1981, 10.1126/science.7466396

81. I. Couzin and J. Krause, Self-organization and collective behavior in vertebrates, Advances in the Study of Behavior 32: 1–75, 2003, 10.1016/S0065-3454(03)01001-5

82. D. Sumpter, Collective Animal Behavior, Princeton University Press, 2010

83. D. G. Rand and M. A. Nowak, Human cooperation, Trends in Cognitive Sciences 17: 413–425, 2013, 10.1016/j.tics.2013.06.003

84. C. Hilbe, K. Chatterjee, and M. A. Nowak, Partners and rivals in direct reciprocity, Nature Human Behaviour 2: 469–477, 2018, 10.1038/s41562-018-0320-9

85. T. Johnson, C. T. Dawes, J. H. Fowler, and O. Smirnov, Slowing COVID-19 transmission as a social dilemma: Lessons for government officials from interdisciplinary research on cooperation, Journal of Behavioral Public Administration 3: 1–13, 2020, 10.30636/jbpa.31.150

86. Z. Wu, Social distancing is a social dilemma game played by every individual against his/her population, PLoS ONE 16: e0255543, 2021, 10.1371/journal.pone.0255543

87. G. Lobinska, A. Pauzner, Arne Traulsen, et al., Evolution of resistance to COVID-19 vaccination with dynamic social distancing, Nature Human Behaviour 6: 193–206, 2022, 10.1038/s41562-021-01281-8

88. A. Traulsen, S. A. Levin, and C. M. Saad-Roy, Individual costs and societal benefits of interventions during the COVID-19 pandemic, medRxiv, February 08, 2023, 10.1101/2023.02.08.23285651

89. D. J. Watts and S. H. Strogatz, Collective dynamics of ‘small-world’ networks, Nature 393: 440–442, 1998, 10.1038/30918

90. S. Milgram, The small world problem, Psychology Today 2: 60–67, 1967

91. M. Kochen, (ed.), The Small World, Ablex, Norwood, NJ, 1989

92. J. Guare, Six Degrees of Separation: A Play, Vintage Books, New York, 1990

93. I. D. Couzin, J. Krause, N. R. Franks, and S. A. Levin, Effective leadership and decision-making in animal groups on the move, Nature 433: 513–516, 2005, 10.1038/nature03236

94. J. R. G. Dyer, C. C. Ioannou, L. J. Morrell, et al., Consensus decision making in human crowds, Animal Behaviour 75: 461–470., 2008, 10.1016/j.anbehav.2007.05.010

95. L. Conradt and T. J. Roper, Conflicts of interest and the evolution of decision sharing, Philos Trans R Soc Lond B Biol Sci. 364: 807–819, 2009, 10.1098/rstb.2008.0257

96. D. J. Sumpter and S. C. Pratt, Quorum responses and consensus decision making, Philos Trans R Soc Lond B Biol Sci. 364: 743–753, 2009, 10.1098/rstb.2008.0204.

97. J. R. Dyer, A. Johansson, D. Helbing, et al., Leadership, consensus decision making and collective behaviour in humans, Philos Trans R Soc Lond B Biol Sci. 364: 781–789, 2009, 10.1098/rstb.2008.0233.

98. R. Singh and R. Adhikari, Age-structured impact of social distancing on the COVID-19 epidemic in India, 2003.12055 [q-bio.PE], 2020, 10.48550/arXiv.2003.12055

99. C. Kadelka, Projecting social contact matrices to populations stratified by binary attributes with known homophily, Mathematical Biosciences and Engineering 20: 3282–3300, 2023, 10.3934/mbe.2023154

100. W. H. Sandholm, Population Games and Evolutionary Dynamics, The MIT Press, 2010.

101. Y. Huang, Y. Hao, M. Wang, et al., Optimality and stability of symmetric evolutionary games with applications in genetic selection, Journal of Mathematical Biosciences and Engineering 12: 503–523, 2015, 10.3934/mbe.2015.12.503

